## Supplementary figures and images for "Immunogenicity following two doses of BBIBP-CorV vaccine and a third booster dose with viral vector and Mrna COVID-19 vaccines against delta and omicron variants in prime immunized adults with two doses of BBIBP-CorV vaccine"

### Supplement figure 1

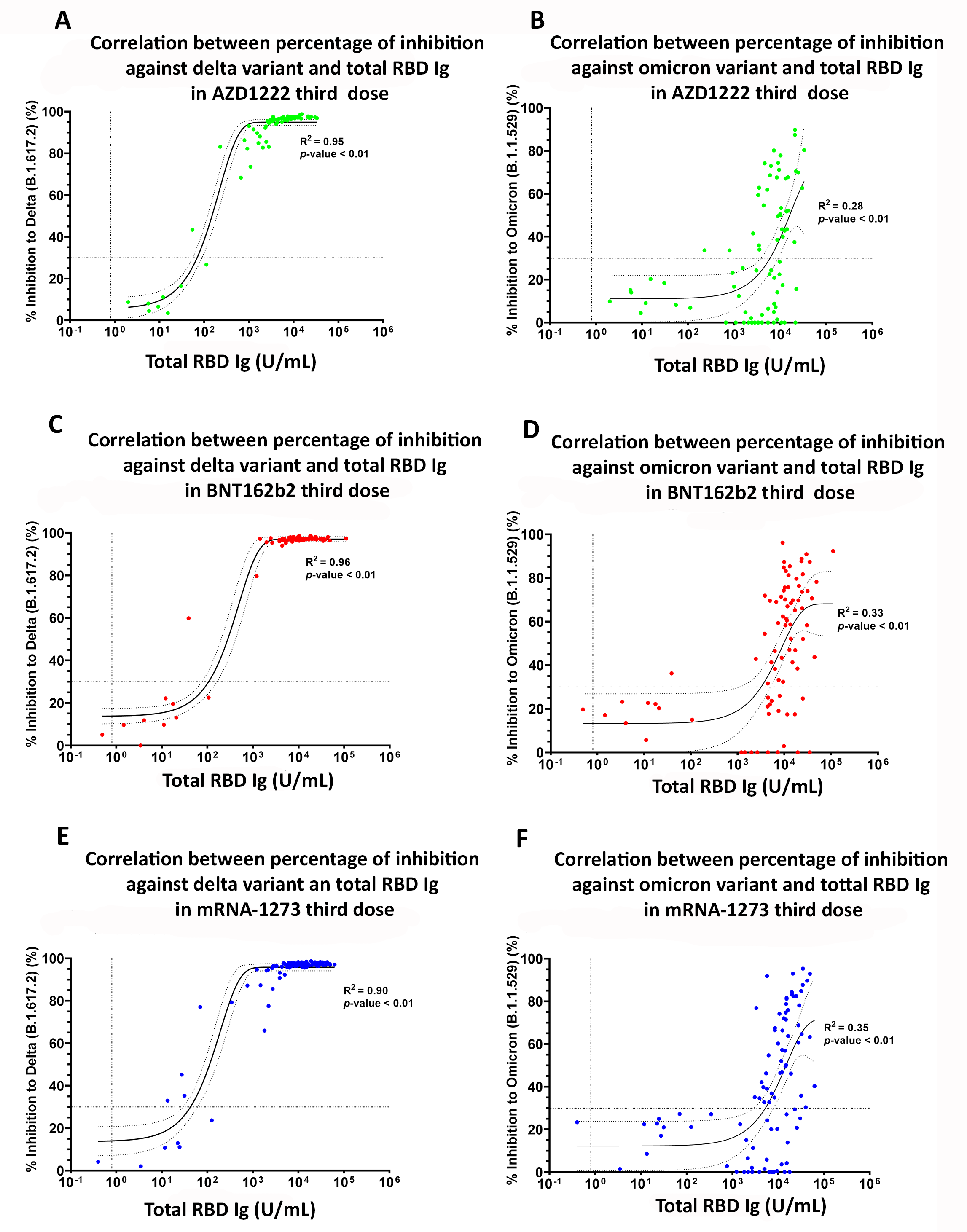

### Supplement figure 2

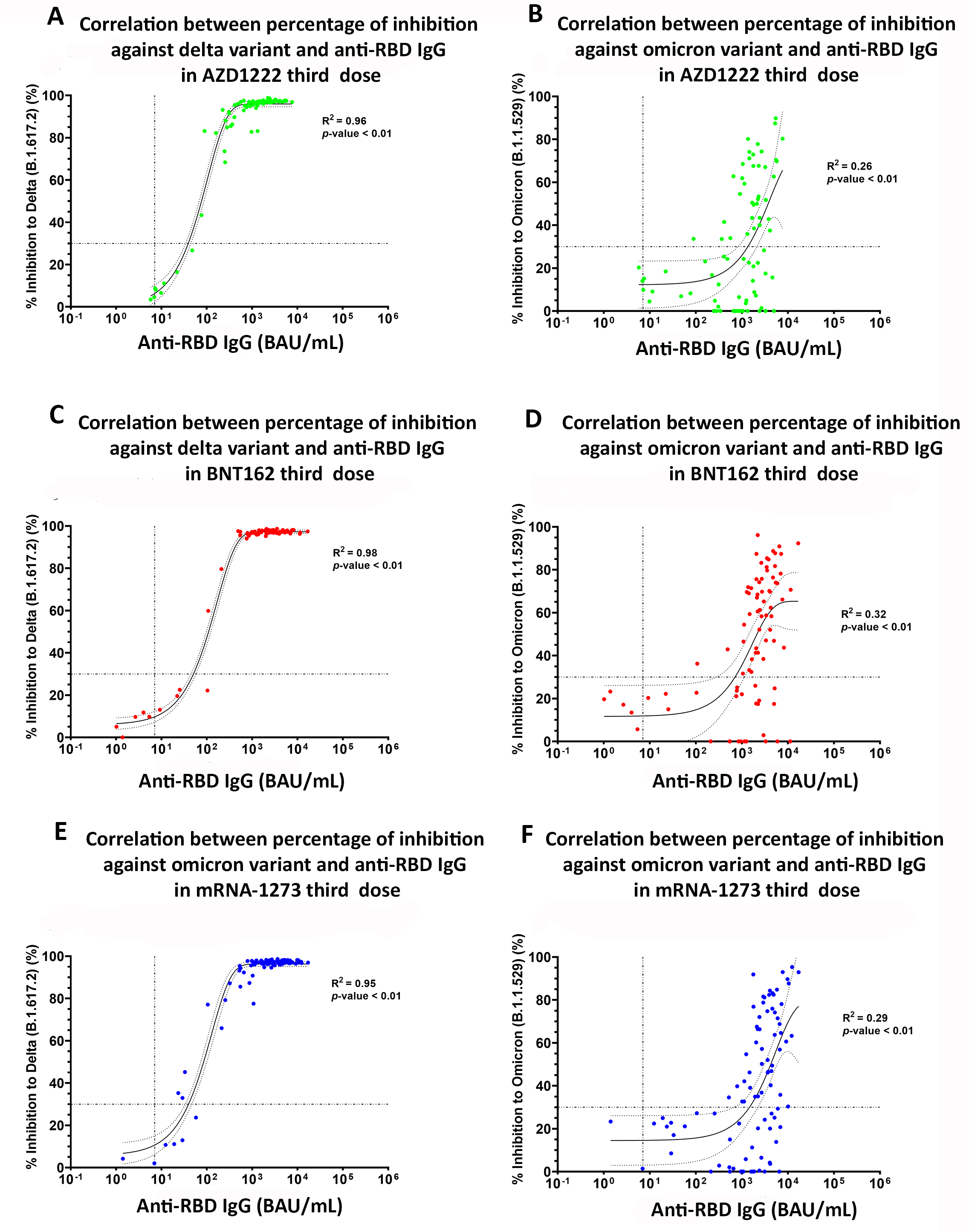
